# Health-Related Quality of Life, Healthcare Utilization, and Expenditure in US Adults With Myocardial Infarction and Heart Failure: The Medical Expenditure Panel Survey 2019 to 2024

**DOI:** 10.64898/2026.09.27.26363737

**Authors:** Iraj Hasan, Zulkifl Jafary, Saim Rana, Harshith Thyagaturu

**Affiliations:** Department of Internal Medicine, West Virginia University, Morgantown, WV, USA; Division of Cardiology, West Virginia University Heart and Vascular Institute, Morgantown, WV, USA

**Author notes:** **Corresponding author**: Iraj Hasan, MD, Department of Internal Medicine, West Virginia University, 1 Medical Center Drive, Morgantown, WV 26506, USA.

**Keywords:** health expenditures, heart failure, myocardial infarction, patient-reported outcome measures, quality of life

## Abstract

**Background:** Myocardial infarction (MI) and heart failure (HF) impose a physical and emotional toll that has not been measured nationally with a standard instrument.

**Methods:** We pooled the 2019 to 2024 Medical Expenditure Panel Survey into 4 mutually exclusive groups of adults aged 18 and older: MI without HF, HF without MI, both conditions, and neither condition. Quality of life was assessed with the Veterans RAND 12-item component summaries, T scores (mean 50, SD 10) on which higher is better and 3 points is minimally important. Survey-weighted linear regression adjusted for demographics, socioeconomic factors, and comorbidity.

**Results:** Among 79,853 adults representing 253.8 million US adults, 4.2% (3,340) had MI alone, 0.6% (480) HF alone, and 0.3% (236) both. Unadjusted physical scores ranged from 33.2 with HF alone to 39.1 with MI alone, versus 50.5 with neither. Adjusted physical scores were lower by 3.84 points (95% CI, 3.20-4.47) with MI alone, 8.32 (95% CI, 7.04-9.61) with HF alone, and 7.74 (95% CI, 5.73-9.76) with both, each beyond the minimally important threshold. Adjusted mental scores were lower by 1.35 points with MI alone, 1.20 with HF alone, and 1.69 with both, all below that threshold. Adults with both conditions did not differ from those with HF alone (0.58 points; 95% CI, -1.76 to 2.92). Emergency visits, inpatient discharges, and expenditure were higher in every cardiac group and highest with both conditions, where annual expenditure reached $37,198 versus $7,845 with neither.

**Conclusions:** Physical quality of life fell steadily across the cardiac groups, with HF rather than prior MI accounting for most of it. The difference in mental quality of life did not persist after adjustment. Utilization and spending were highest with both conditions.

**WHAT IS KNOWN:** Myocardial infarction and heart failure are leading contributors to morbidity and to healthcare spending in the United States, and expenditure for each has been estimated in nationally representative data.

Patient-reported outcomes have been described within broader atherosclerotic cardiovascular disease populations, where depression risk is associated with worse reported health and higher spending.

**WHAT THE STUDY ADDS:** This study quantifies health-related quality of life against the general US adult population for myocardial infarction and heart failure held apart from one another, and shows that the physical burden of heart failure is more than twice that of prior infarction.

Carrying both conditions adds no measurable decrement beyond heart failure alone, and the difference in mental quality of life does not persist after adjustment for age and coexisting illness.

Unadjusted annual spending rose from $19,876 in myocardial infarction alone to $34,002 in heart failure alone and $37,198 with both conditions, against $7,845 in adults with neither. Spending was highest among adults with both conditions, while physical quality of life was lowest among those with heart failure alone. Expenditure alone may therefore not capture how patients in these groups report feeling.

## INTRODUCTION

Cardiovascular disease is the leading source of death and healthcare spending in the United States, and its burden is projected to grow. Heart failure prevalence among US adults is expected to rise from 2.7% in 2020 to 3.8% by 2050, with coronary disease rising from 7.8% to 9.2% over the same period.^1^ Heart failure alone accounts for an estimated $179.5 billion in annual expenditure, and spending on the condition is growing faster than spending on myocardial infarction, diabetes, or cancer.^2^ Both conditions fall disproportionately on older adults and on those with limited income and public insurance.

Medical expenditures in these conditions are concentrated in hospital care. A diagnosis of heart failure has been associated with $3,594 in annual incremental expenditure after adjustment for demographics and comorbidities.^2^ Acute myocardial infarction has been associated with $7,076 in excess direct medical expenditure.^3^ Coexisting illness adds further cost. Among hypertensive adults, congestive heart failure carried the largest per-person expenditure effect of the 15 most frequent coexisting conditions, at $7,380.^4^ Out-of-pocket burden in heart failure has in turn been linked to cost-related medication non-adherence, which affected 7.9% of patients in a recent national analysis.^5^

Expenditure captures only part of what these conditions cost. Myocardial infarction and heart failure impose an emotional toll as well as a physical one. Patients live with exertional limitation, breathlessness, fear of recurrence, and uncertainty about prognosis, and none of that is measured by spending. Health-related quality of life instruments are built to capture both dimensions, returning a physical and a mental summary score from the same questionnaire. Patient-reported outcomes have been described among adults with atherosclerotic cardiovascular disease, where women reported poorer patient experience and lower quality of life than men, and where optimal shared decision-making was associated with better perceived health.^6,7^ Depression risk in the same population has been associated with worse reported health and with higher overall and out-of-pocket expenditure.^8^ The component summary scores of the Veterans RAND 12-item Health Survey have been used to quantify this kind of burden in other conditions in the current era of the survey.^9^

The size of that burden has not been quantified in a nationally representative sample. Estimates exist for the expenditure side of both conditions, and patient experience has been characterized within broader cardiovascular populations. Adults living with myocardial infarction or heart failure have not been measured against the general adult population on a standard quality of life instrument. The physical and emotional components of that burden have not been compared with one another, and it is not known whether carrying both conditions compounds it. It is also unknown whether quality of life in these populations shifted across a period that included the COVID-19 pandemic.

To address these gaps, we used six years of the Medical Expenditure Panel Survey to 1) quantify health-related quality of life among US adults with myocardial infarction alone, heart failure alone, both conditions, and neither, 2) determine whether carrying both conditions adds further burden beyond either alone, and 3) describe healthcare utilization and expenditure across the same groups. We first describe the characteristics of the four cohorts. We then compare unadjusted and adjusted component summary scores across them, contrast the combined cohort against heart failure alone, report healthcare utilization and expenditure, and examine whether quality of life changed between 2019 and 2024.

## METHODS

### Data Sources

The Medical Expenditure Panel Survey has been fielded annually since 1996 under the Agency for Healthcare Research and Quality. It draws on three linked sources, households, employers, and the clinicians, hospitals, and pharmacies that treat and supply them, and is the primary national source for linked data on health service use and spending. Households enter the sample from respondents to the previous year National Health Interview Survey. Each household is interviewed on five occasions over two calendar years. Because the released files are deidentified and publicly available, institutional review board approval was not required for this analysis.

### Study Participants

Six survey years, 2019 through 2024, were combined for this analysis. Eligibility was confined to adults aged 18 years and older who completed the self-administered questionnaire and had a scored VR-12, because respondents without those data carry no quality of life measure and are assigned a questionnaire weight of zero. That restriction left 79,853 respondents, representing 253.8 million US adults. Both the Full-Year Consolidated files and the Medical Conditions files were used. Myocardial infarction was taken from MIDX, a priority condition variable recording whether a physician had ever told the respondent they had a heart attack. Heart failure was taken from the Medical Conditions files, where a respondent counted as affected if any condition record for that survey year carried Clinical Classification Software Refined code CIR019, matched to the person by DUPERSID. Cross-tabulating the two conditions produced 4 non-overlapping groups: (a) myocardial infarction without heart failure (MI+/HF-, n=3,340); (b) heart failure without myocardial infarction (MI-/HF+, n=480); (c) both conditions (MI+/HF+, n=236); and (d) neither condition (MI-/HF-, n=75,797). Pooling six years rather than one was done to build sufficient numbers in the two smaller groups.

### Measurements

Three measures were drawn from the files. Quality of life was the outcome entered into regression models; resource use and expenditure were summarized descriptively and were not modeled.

Quality of life came from the VR-12, a 12-item instrument adapted from the SF-36. It departs from the SF-36 and SF-12 by scoring the role-physical and role-emotional domains on a graded five-level response rather than a yes-or-no answer, which reduced clustering at the extremes of the scale. Published crosswalks allow VR-12 and SF-36 results to be compared. Responses are scored centrally by the survey and released as physical and mental component summaries, VPCS42 and VMCS42. Each is normed so that the US population averages 50 with a standard deviation of 10, and a higher value denotes better reported health. A respondent scoring 40 therefore sits a full standard deviation below the national average. Three points is the accepted minimally important difference and is applied throughout to separate statistical significance from clinical relevance. Reserved codes for inapplicable responses were set to missing before estimation. Psychological distress was captured by the Kessler-6 scale and depressive symptoms by the two-item Patient Health Questionnaire.

Seven counts described resource use over the year of observation: visits to office-based providers, visits to hospital outpatient departments, emergency department visits, inpatient discharges, nights spent in hospital, prescriptions filled, and days of home health care.

Expenditure in this survey means money actually paid for care delivered during the calendar year, counting what the family paid itself alongside payments made by private insurers, Medicare, Medicaid, and any other payer. Household accounts of service use are verified against records obtained from the treating hospitals, clinicians, and pharmacies before the figures are finalized in the Full-Year Consolidated files. Spending was examined in total and by service line, covering office-based care, hospital outpatient care, emergency care, inpatient stays, prescription medicines, home health care, dental care, and other equipment and services. Out-of-pocket spending was the portion the family paid directly.

Adjustment variables comprised age, sex, race and ethnicity, marital status, education, family income relative to the federal poverty threshold, insurance category, and census region. Coexisting illness comprised hypertension, hyperlipidemia, diabetes, cancer, arthritis, asthma, and emphysema. Sex and race and ethnicity were adjustment covariates, not tested for effect modification of the cardiac group differences.

### Statistical Analysis

Group differences in baseline characteristics, coexisting illness, quality of life, resource use, and spending were tested with the design-based Rao-Scott F test for categorical variables and the adjusted Wald test for continuous variables. Because six years are pooled, the questionnaire weight for each respondent was divided by 6 so the combined file represents a single year of the US population. Variance estimation used Taylor series linearization with the survey strata and primary sampling units, and every group-specific estimate used subpopulation commands to preserve the full sample design in the standard errors.

Differences in component scores across the 4 groups were estimated by linear regression fitted in three stages. The first stage held only age category and sex constant; the second added race and ethnicity, education, income category, insurance category, marital status, census region, and survey year; and the third, which supplies the estimates reported as primary, further introduced hypertension, hyperlipidemia, and a count of non-cardiac coexisting conditions. Coefficients are expressed as the number of T-score points separating each cardiac group from adults free of both conditions. The combined group was additionally compared against the heart failure group within the fully adjusted specification. Change over time was assessed by regressing each component score on calendar year separately within each group. Resource use and spending appear as unadjusted weighted means only. Any estimate resting on fewer than 60 unweighted observations is marked and left uninterpreted. Analyses were carried out in Stata 18 (StataCorp LLC, College Station, TX), with two-sided P<0.05 taken as the threshold for significance.

Iraj Hasan and Zulkifl Jafary had full access to the data and take responsibility for its integrity and the analysis.

## RESULTS

### Sample Characteristics

Table 1 shows the characteristics of the four groups. Out of 79,853 adults in the six year pooled data, 4.2% (3,340) had myocardial infarction alone, 0.6% (480) had heart failure alone, and 0.3% (236) had both conditions. The remaining 94.9% (75,797) had neither. Mean age was 67.4 years in patients with myocardial infarction alone, 67.5 years in patients with heart failure alone, and 68.6 years in patients with both conditions, against 47.7 years in patients with neither condition. Patients with myocardial infarction alone were 64.2% male and patients with heart failure alone were 59.2% female. Hypertension was present in 81.2% of patients with myocardial infarction alone, 83.7% of patients with heart failure alone, and 81.1% of patients with both conditions, against 31.2% of patients with neither condition. Diabetes was present in 36.8% of patients with myocardial infarction alone, 35.0% of patients with heart failure alone, and 42.8% of patients with both conditions, against 10.6% of patients with neither condition. The proportion uninsured at any time in the year was 0.2% in patients with heart failure alone and 0.4% in patients with both conditions, against 7.3% in patients with neither condition. Serious psychological distress and depression screens were more frequent in every cardiac group.

**Table 1.**
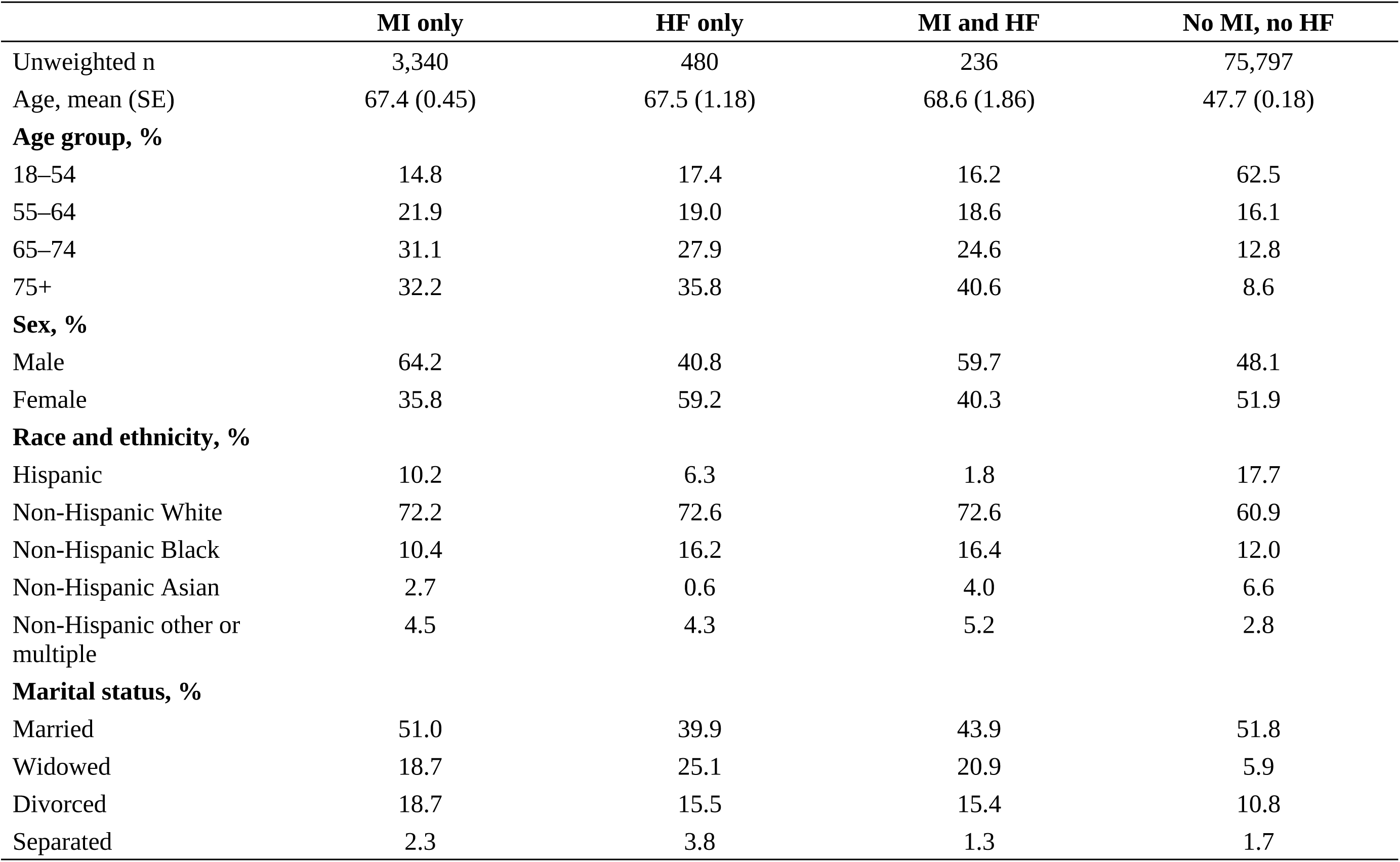
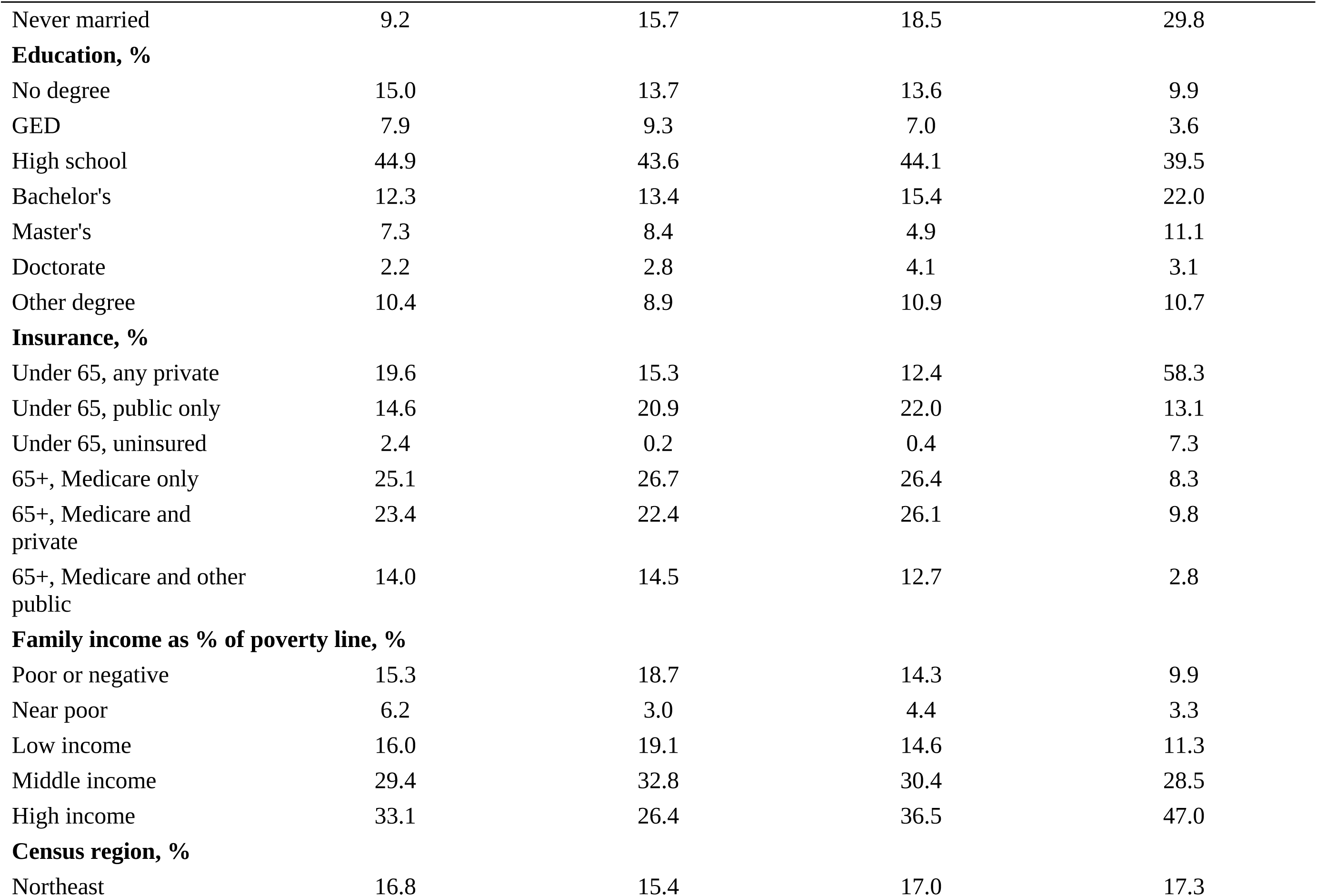
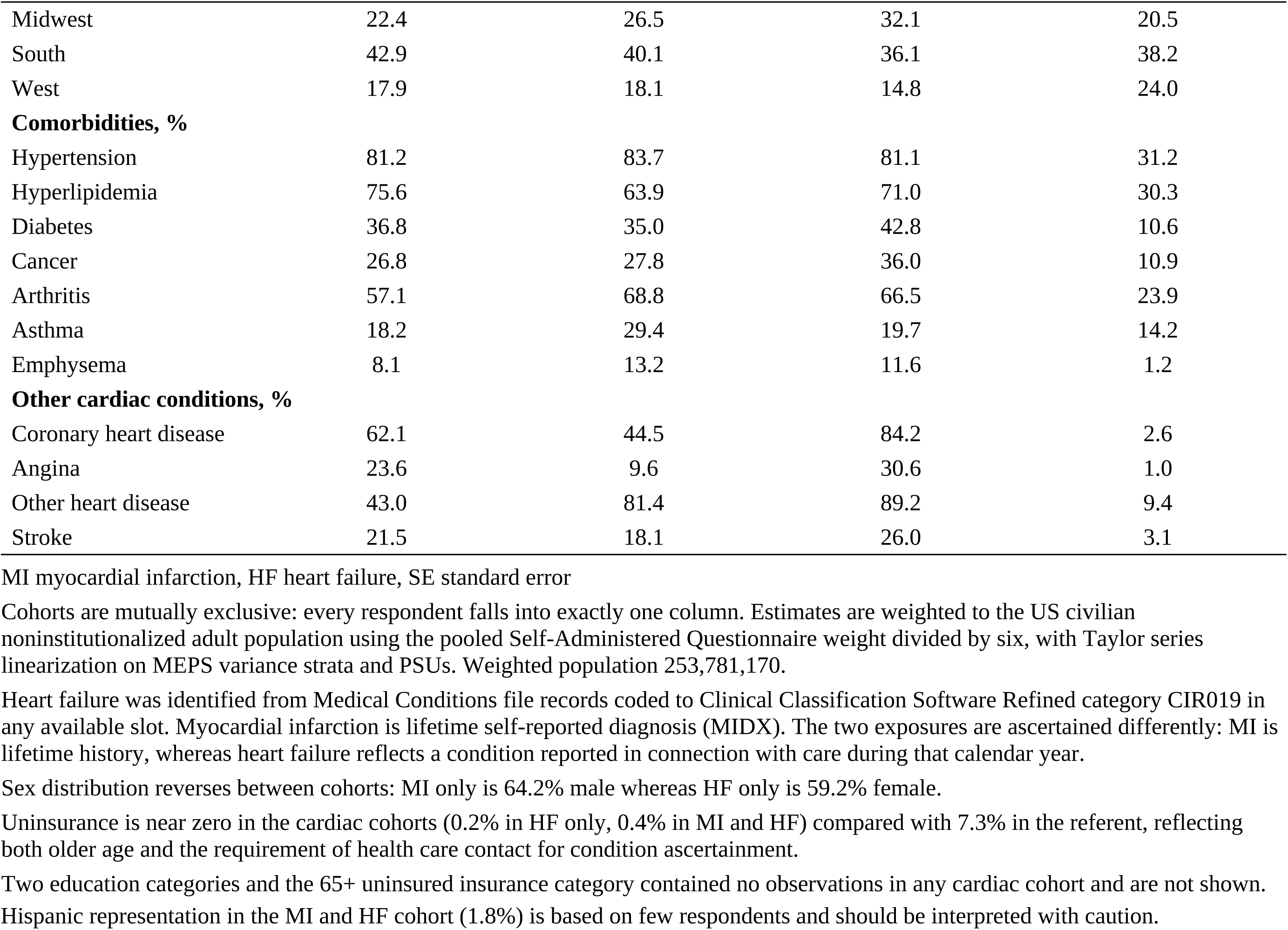
Demographic and baseline characteristics.

|  | MI only | HF only | MI and HF | No MI, no HF |
| --- | --- | --- | --- | --- |
| Unweighted n | 3,340 | 480 | 236 | 75,797 |
| Age, mean (SE) | 67.4 (0.45) | 67.5 (1.18) | 68.6 (1.86) | 47.7 (0.18) |
| **Age group, %** |  |  |  |  |
| 18–54 | 14.8 | 17.4 | 16.2 | 62.5 |
| 55–64 | 21.9 | 19.0 | 18.6 | 16.1 |
| 65–74 | 31.1 | 27.9 | 24.6 | 12.8 |
| 75+ | 32.2 | 35.8 | 40.6 | 8.6 |
| **Sex, %** |  |  |  |  |
| Male | 64.2 | 40.8 | 59.7 | 48.1 |
| Female | 35.8 | 59.2 | 40.3 | 51.9 |
| **Race and ethnicity, %** |  |  |  |  |
| Hispanic | 10.2 | 6.3 | 1.8 | 17.7 |
| Non-Hispanic White | 72.2 | 72.6 | 72.6 | 60.9 |
| Non-Hispanic Black | 10.4 | 16.2 | 16.4 | 12.0 |
| Non-Hispanic Asian | 2.7 | 0.6 | 4.0 | 6.6 |
| Non-Hispanic other or multiple | 4.5 | 4.3 | 5.2 | 2.8 |
| **Marital status, %** |  |  |  |  |
| Married | 51.0 | 39.9 | 43.9 | 51.8 |
| Widowed | 18.7 | 25.1 | 20.9 | 5.9 |
| Divorced | 18.7 | 15.5 | 15.4 | 10.8 |
| Separated | 2.3 | 3.8 | 1.3 | 1.7 |
| Never married | 9.2 | 15.7 | 18.5 | 29.8 |
| **Education, %** |  |  |  |  |
| No degree | 15.0 | 13.7 | 13.6 | 9.9 |
| GED | 7.9 | 9.3 | 7.0 | 3.6 |
| High school | 44.9 | 43.6 | 44.1 | 39.5 |
| Bachelor's | 12.3 | 13.4 | 15.4 | 22.0 |
| Master's | 7.3 | 8.4 | 4.9 | 11.1 |
| Doctorate | 2.2 | 2.8 | 4.1 | 3.1 |
| Other degree | 10.4 | 8.9 | 10.9 | 10.7 |
| **Insurance, %** |  |  |  |  |
| Under 65, any private | 19.6 | 15.3 | 12.4 | 58.3 |
| Under 65, public only | 14.6 | 20.9 | 22.0 | 13.1 |
| Under 65, uninsured | 2.4 | 0.2 | 0.4 | 7.3 |
| 65+, Medicare only | 25.1 | 26.7 | 26.4 | 8.3 |
| 65+, Medicare and private | 23.4 | 22.4 | 26.1 | 9.8 |
| 65+, Medicare and other public | 14.0 | 14.5 | 12.7 | 2.8 |
| **Family income as % of poverty line, %** |  |  |  |  |
| Poor or negative | 15.3 | 18.7 | 14.3 | 9.9 |
| Near poor | 6.2 | 3.0 | 4.4 | 3.3 |
| Low income | 16.0 | 19.1 | 14.6 | 11.3 |
| Middle income | 29.4 | 32.8 | 30.4 | 28.5 |
| High income | 33.1 | 26.4 | 36.5 | 47.0 |
| **Census region, %** |  |  |  |  |
| Northeast | 16.8 | 15.4 | 17.0 | 17.3 |
| Midwest | 22.4 | 26.5 | 32.1 | 20.5 |
| South | 42.9 | 40.1 | 36.1 | 38.2 |
| West | 17.9 | 18.1 | 14.8 | 24.0 |
| **Comorbidities, %** |  |  |  |  |
| Hypertension | 81.2 | 83.7 | 81.1 | 31.2 |
| Hyperlipidemia | 75.6 | 63.9 | 71.0 | 30.3 |
| Diabetes | 36.8 | 35.0 | 42.8 | 10.6 |
| Cancer | 26.8 | 27.8 | 36.0 | 10.9 |
| Arthritis | 57.1 | 68.8 | 66.5 | 23.9 |
| Asthma | 18.2 | 29.4 | 19.7 | 14.2 |
| Emphysema | 8.1 | 13.2 | 11.6 | 1.2 |
| **Other cardiac conditions, %** |  |  |  |  |
| Coronary heart disease | 62.1 | 44.5 | 84.2 | 2.6 |
| Angina | 23.6 | 9.6 | 30.6 | 1.0 |
| Other heart disease | 43.0 | 81.4 | 89.2 | 9.4 |
| Stroke | 21.5 | 18.1 | 26.0 | 3.1 |

MI myocardial infarction, HF heart failure, SE standard error
Cohorts are mutually exclusive: every respondent falls into exactly one column. Estimates are weighted to the US civilian noninstitutionalized adult population using the pooled Self-Administered Questionnaire weight divided by six, with Taylor series linearization on MEPS variance strata and PSUs. Weighted population 253,781,170.
Heart failure was identified from Medical Conditions file records coded to Clinical Classification Software Refined category CIR019 in any available slot. Myocardial infarction is lifetime self-reported diagnosis (MIDX). The two exposures are ascertained differently: MI is lifetime history, whereas heart failure reflects a condition reported in connection with care during that calendar year.
Sex distribution reverses between cohorts: MI only is 64.2% male whereas HF only is 59.2% female.
Uninsurance is near zero in the cardiac cohorts (0.2% in HF only, 0.4% in MI and HF) compared with 7.3% in the referent, reflecting both older age and the requirement of health care contact for condition ascertainment.
Two education categories and the 65+ uninsured insurance category contained no observations in any cardiac cohort and are not shown.
Hispanic representation in the MI and HF cohort (1.8%) is based on few respondents and should be interpreted with caution.

### Health-Related Quality of Life

Table 2 shows unadjusted component summary scores by group. Higher scores indicate better health, and every score below 50 sits under the US general population average. The mean Physical Component Summary was 39.1 in patients with myocardial infarction alone, 33.2 in patients with heart failure alone, and 34.1 in patients with both conditions, against 50.5 in patients with neither condition. Each cardiac group therefore scored worse than patients with neither condition, and patients with heart failure alone scored worst. Those raw gaps range from 11.4 to 17.3 points, between three and six times the minimally important difference, and are large enough for a patient to notice. The mean Mental Component Summary was 49.9 in patients with myocardial infarction alone, 48.9 in patients with heart failure alone, and 49.2 in patients with both conditions, against 51.5 in patients with neither condition. These scores also fall below patients with neither condition, but the raw gaps of 1.6 to 2.6 points are smaller than the minimally important difference before any adjustment.

**Table 2.** Health-related quality of life, medical resource utilization, and medical expenditures.

|  | MI only | HF only | MI and HF | No MI, no HF |
| --- | --- | --- | --- | --- |
| Unweighted n | 3,340 | 480 | 236 | 75,797 |
| **Health-related quality of life, mean (SE)** |  |  |  |  |
| VR-12 Physical Component Summary | 39.1 (0.35) | 33.2 (0.80) | 34.1 (1.01) | 50.5 (0.10) |
| VR-12 Mental Component Summary | 49.9 (0.30) | 48.9 (0.69) | 49.2 (0.89) | 51.5 (0.08) |
| Kessler-6 distress score | 4.10 (0.16) | 4.79 (0.34) | 4.55 (0.45) | 3.08 (0.04) |
| PHQ-2 depression score | 1.02 (0.05) | 1.25 (0.11) | 1.20 (0.13) | 0.64 (0.01) |
| **Medical resource utilization, mean** |  |  |  |  |
| Office-based visits | 12.24 | 17.88 | 17.03 | 7.58 |
| Outpatient visits | 2.59 | 3.85 | 3.77 | 1.03 |
| Emergency visits | 0.57 | 0.80 | 1.02 | 0.19 |
| Inpatient discharges | 0.33 | 0.61 | 0.77 | 0.08 |
| Nights in hospital | 1.76 | 3.98 | 4.80 | 0.35 |
| Prescription fills | 33.31 | 46.38 | 42.76 | 10.63 |
| Home health days | 12.68 | 19.87 | 21.37 | 2.72 |
| **Healthcare expenditures, mean \$** | | | | |
| Office-based | 3,974 | 5,742 | 7,899 | 2,171 |
| Outpatient | 1,690 | 2,946 | 2,167 | 979 |
| Emergency | 517 | 502 | 1,919 | 243 |
| Inpatient | 5,265 | 10,562 | 14,135 | 1,402 |
| Prescription drugs | 5,959 | 8,850 | 7,076 | 1,977 |
| Home health | 1,222 | 3,380 | 2,462 | 277 |
| Other equipment and services | 596 | 1,085 | 766 | 243 |
| Out-of-pocket | 1,594 | 2,480 | 3,631 | 1,072 |
| Total expenditure | 19,876 | 34,002 | 37,198 | 7,845 |

MI myocardial infarction, HF heart failure, HRQoL health-related quality of life, PHQ-2 Patient Health Questionnaire-2, SE standard error
VR-12 component summaries are norm-based T-scores with a US population mean of 50 and standard deviation of 10; higher scores indicate better health-related quality of life. A difference of 3 points is conventionally regarded as the minimally important difference. Scores are taken from the MEPS-supplied VR-12 summary variables (VPCS42, VMCS42). Reserved codes for inapplicable (−1) and not computable (−15) were set to missing before estimation.
Expenditures are nominal dollars, not adjusted for inflation across the 2019–2024 period. All estimates are weighted using the pooled Self-Administered Questionnaire weight.
Unadjusted physical component decrements relative to the referent are 11.4 points for MI only, 17.2 for HF only, and 16.4 for MI and HF, all well beyond the minimally important difference. Corresponding mental component decrements are 1.6, 2.6, and 2.3 points, all below it.
The MI and HF cohort does not differ meaningfully from HF only on either component (physical 34.1 versus 33.2; confidence intervals overlap substantially), indicating that prior infarction adds little once heart failure is present.
Mean emergency expenditure in the MI and HF cohort (\$1,919) has a 95% confidence interval spanning zero (−\$581 to \$4,418) and is driven by a small number of high-cost respondents.
These estimates are unadjusted. The referent cohort is on average 20 years younger than the cardiac cohorts, so a substantial portion of these differences reflects age and comorbidity rather than cardiac disease.

### Adjusted Differences in Quality of Life

Table 5 and Figure 1 show the results of the adjusted models. All estimates below come from Model 3, adjusted for sociodemographic factors, coexisting conditions, and survey year. Negative values indicate worse health than patients with neither condition. Patients with myocardial infarction alone had a lower Physical Component Summary score [-3.84 points (95% CI -4.47 to -3.20, P<0.001)]. Patients with heart failure alone had a larger reduction [-8.32 points (95% CI -9.61 to -7.04, P<0.001)]. Patients with both conditions had a reduction of similar size [-7.74 points (95% CI -9.76 to -5.73, P<0.001)]. All 3 exceeded the 3-point minimally important difference, and each represents a decrement a patient would be expected to notice.

**Figure 1.**
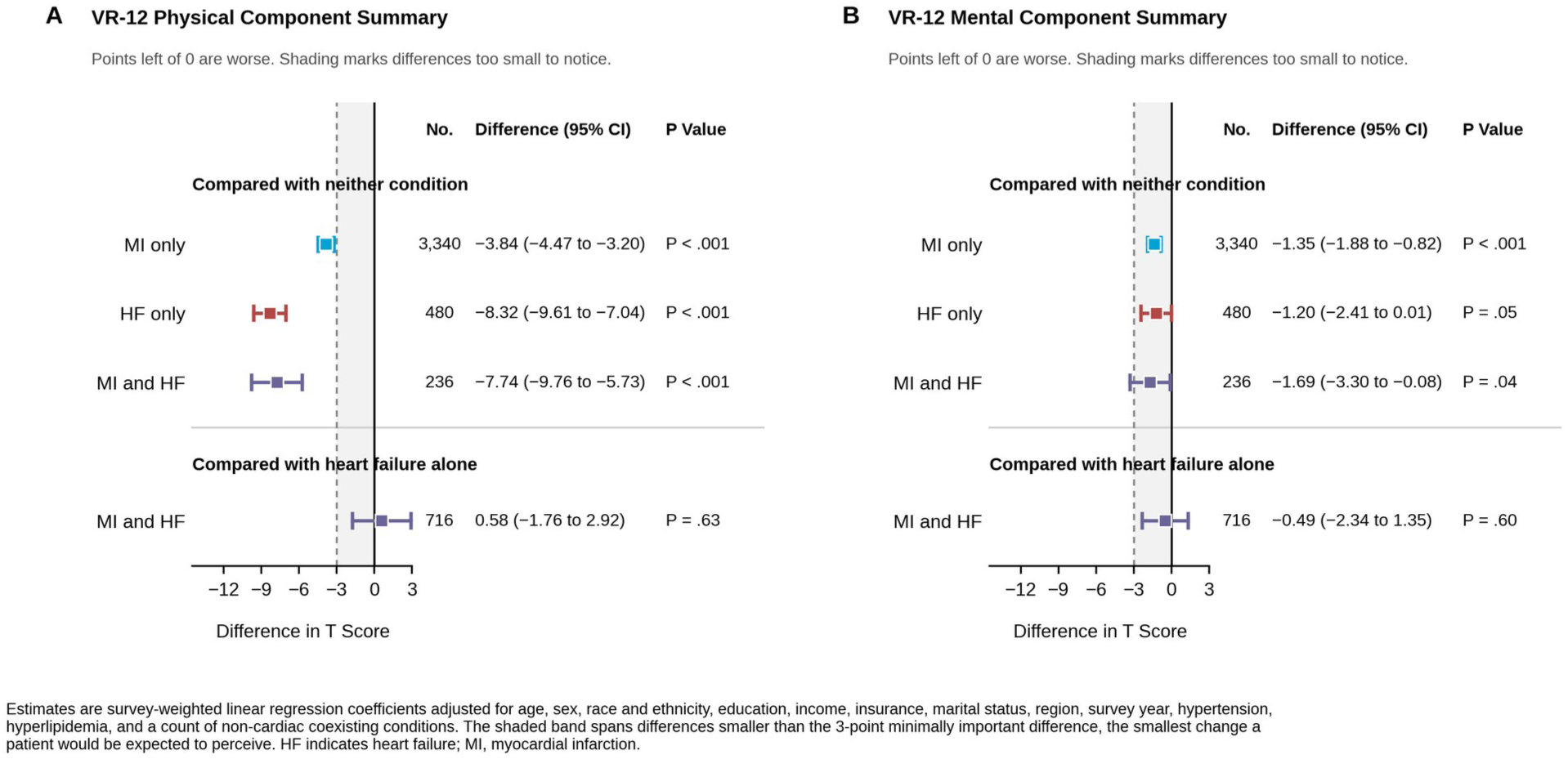
Adjusted differences in quality of life by cohort. A, Physical Component Summary. B, Mental Component Summary. Each panel has two blocks. The upper block compares each cardiac cohort with adults having neither condition. The lower block compares adults carrying both conditions with those having heart failure alone. Markers are adjusted differences and bars are 95% confidence intervals. Shading spans differences below the 3-point minimally important difference.

Differences in mental quality of life were smaller. Patients with myocardial infarction alone had a lower Mental Component Summary score [-1.35 points (95% CI -1.88 to -0.82, P<0.001)]. The reduction in patients with heart failure alone did not reach significance [-1.20 points (95% CI -2.41 to 0.01, P=0.052)]. Patients with both conditions had a small reduction [-1.69 points (95% CI -3.30 to -0.08, P=0.039)]. None of the 3 reached the 3-point minimally important difference, and none represents a decrement a patient would be expected to notice.

Figure S1 shows the estimates across the three nested models. Adjustment reduced every difference. Differences in physical quality of life fell from 7.49 to 3.84 points in patients with myocardial infarction alone, from 12.99 to 8.32 points in patients with heart failure alone, and from 12.08 to 7.74 points in patients with both conditions. Differences in mental quality of life fell from 3.04 to 1.35 points in patients with myocardial infarction alone, from 3.54 to 1.20 points in patients with heart failure alone, and from 3.64 to 1.69 points in patients with both conditions. Every mental estimate crossed below the minimally important difference only after adjustment for coexisting conditions.

### Combined Disease Versus Heart Failure Alone

Patients with both conditions did not differ from patients with heart failure alone on either component in the fully adjusted model. The difference in physical quality of life was not significant [0.58 points (95% CI -1.76 to 2.92, P=0.626)]. The difference in mental quality of life was also not significant [-0.49 points (95% CI -2.34 to 1.35, P=0.599)]. Both confidence intervals cross zero. The same null was present before adjustment.

### Health Resource Utilization and Expenditures

Table 2 also reports unadjusted utilization, and every measure was higher in the cardiac groups than in patients with neither condition. Office-based visits averaged 12.2 with myocardial infarction alone, 17.9 with heart failure alone, 17.0 with both conditions, and 7.6 with neither. Hospital outpatient visits averaged 2.6 with myocardial infarction alone, 3.9 with heart failure alone, 3.8 with both conditions, and 1.0 with neither. Emergency department visits averaged 0.57 with myocardial infarction alone, 0.80 with heart failure alone, 1.02 with both conditions, and 0.19 with neither. Inpatient discharges averaged 0.33 with myocardial infarction alone, 0.61 with heart failure alone, 0.77 with both conditions, and 0.08 with neither. Nights in hospital averaged 1.76 with myocardial infarction alone, 3.98 with heart failure alone, 4.80 with both conditions, and 0.35 with neither. Prescription fills averaged 33.3 with myocardial infarction alone, 46.4 with heart failure alone, 42.8 with both conditions, and 10.6 with neither, and home health days averaged 12.7 with myocardial infarction alone, 19.9 with heart failure alone, 21.4 with both conditions, and 2.7 with neither. Patients with both conditions recorded the highest use of emergency, inpatient, and home health services.

Table 2 and Figure 2 show unadjusted expenditure. Mean total annual expenditure per person was $19,876 in patients with myocardial infarction alone, $34,002 in patients with heart failure alone, and $37,198 in patients with both conditions, against $7,845 in patients with neither condition. Inpatient care was the largest single component in both heart failure groups, at $10,562 and $14,135. In patients with myocardial infarction alone, prescription drugs at $5,959 exceeded inpatient care at $5,265. Out-of-pocket expenditure was $1,594 in patients with myocardial infarction alone, $2,480 in patients with heart failure alone, and $3,631 in patients with both conditions, against $1,072 in patients with neither condition.

**Figure 2.**
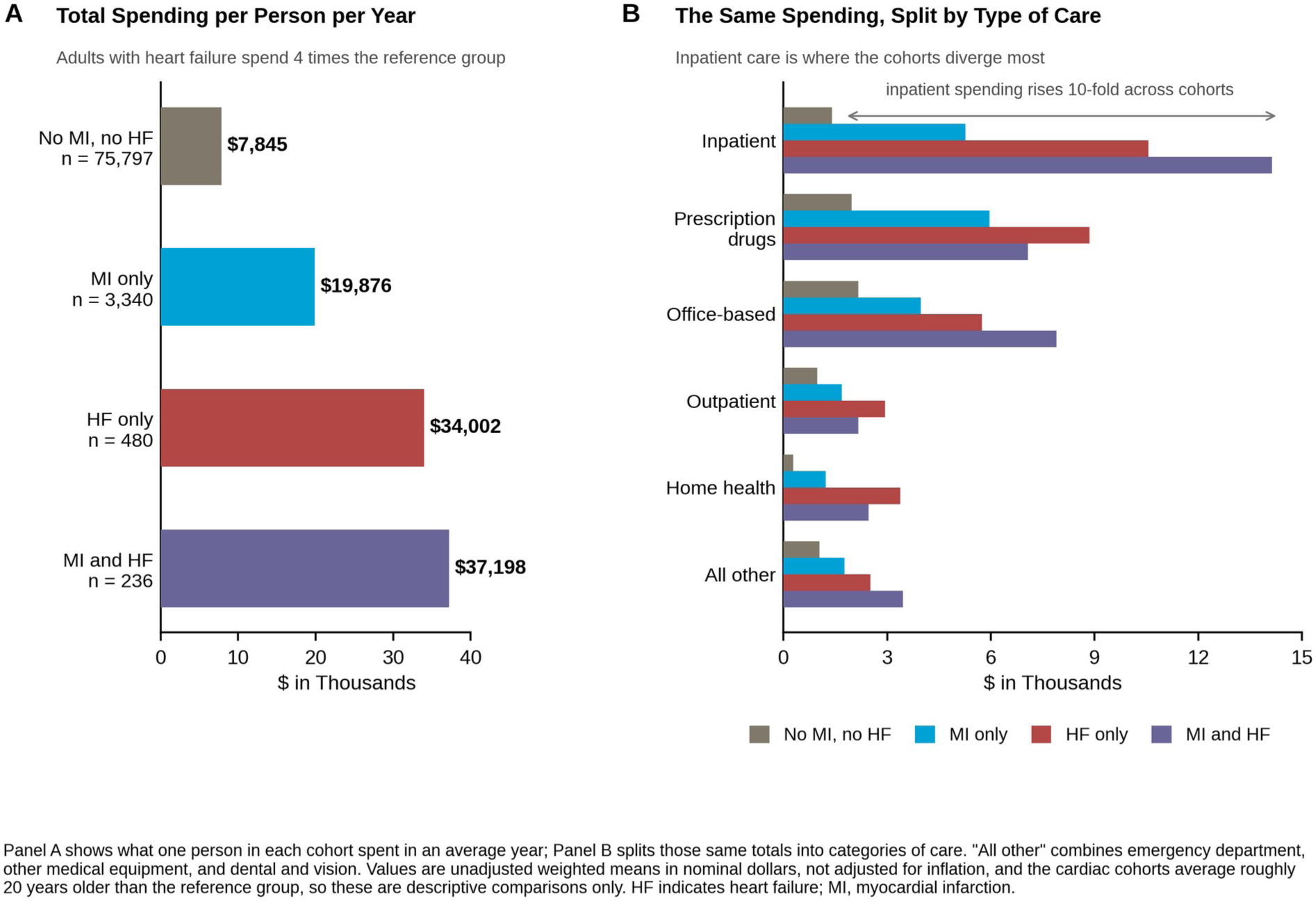
Annual healthcare expenditure per person by cohort. A, Total expenditure per person per year. B, The same totals split by category of care. Values are unadjusted weighted means in nominal dollars.

Figure S2 plots the physical component score against total expenditure for each group. Patients with both conditions recorded the highest expenditure at $37,198 per person, and patients with heart failure alone recorded the lowest physical score at 33.2 points.

### Trends Across the Study Period

Tables 3 and 4 and Figure S3 show component summary scores by calendar year for the two single-condition groups. A trend test, a linear regression of the score on calendar year, asks whether scores rose or fell steadily from 2019 to 2024 rather than moving randomly. Neither group showed a significant linear trend in either component. In patients with myocardial infarction alone, the trend test returned P=0.287 for the physical component and P=0.473 for the mental component. In patients with heart failure alone, the trend test returned P=0.821 for the physical component and P=0.166 for the mental component. Point estimates were stable through 2020. The trend test for patients with both conditions is reported in Table S1 for completeness only. At 23 to 57 observations per year that analysis is underpowered to detect a linear trend, and the result should not be read as evidence of either stability or change.

**Table 3.**
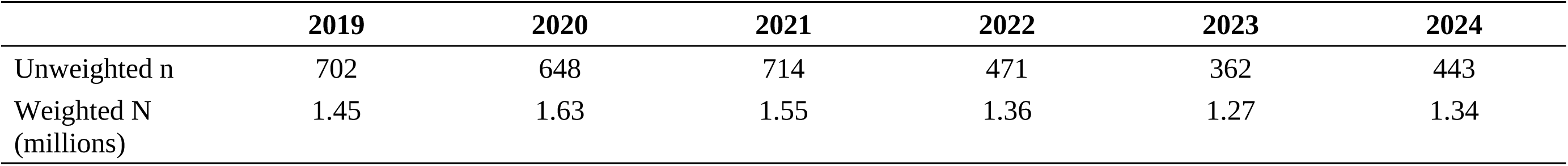
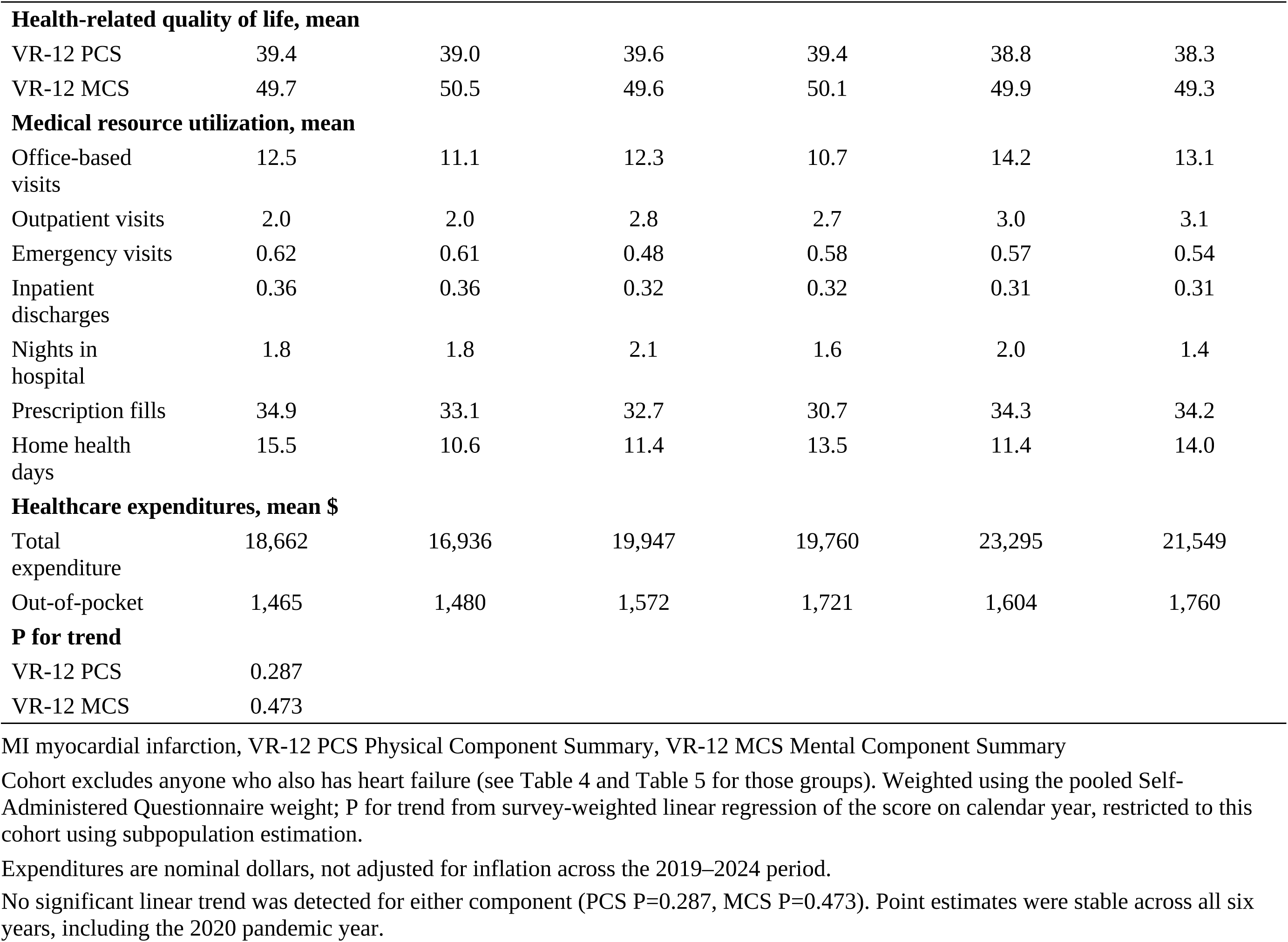
Health-related quality of life, medical resource utilization, and medical expenditures among adults with myocardial infarction only, by year.

|  | 2019 | 2020 | 2021 | 2022 | 2023 | 2024 |
| --- | --- | --- | --- | --- | --- | --- |
| Unweighted n | 702 | 648 | 714 | 471 | 362 | 443 |
| Weighted N (millions) | 1.45 | 1.63 | 1.55 | 1.36 | 1.27 | 1.34 |
| **Health-related quality of life, mean** |  |  |  |  |  |  |
| VR-12 PCS | 39.4 | 39.0 | 39.6 | 39.4 | 38.8 | 38.3 |
| VR-12 MCS | 49.7 | 50.5 | 49.6 | 50.1 | 49.9 | 49.3 |
| **Medical resource utilization, mean** |  |  |  |  |  |  |
| Office-based visits | 12.5 | 11.1 | 12.3 | 10.7 | 14.2 | 13.1 |
| Outpatient visits | 2.0 | 2.0 | 2.8 | 2.7 | 3.0 | 3.1 |
| Emergency visits | 0.62 | 0.61 | 0.48 | 0.58 | 0.57 | 0.54 |
| Inpatient discharges | 0.36 | 0.36 | 0.32 | 0.32 | 0.31 | 0.31 |
| Nights in hospital | 1.8 | 1.8 | 2.1 | 1.6 | 2.0 | 1.4 |
| Prescription fills | 34.9 | 33.1 | 32.7 | 30.7 | 34.3 | 34.2 |
| Home health days | 15.5 | 10.6 | 11.4 | 13.5 | 11.4 | 14.0 |
| **Healthcare expenditures, mean \$** | | | | | | |
| Total expenditure | 18,662 | 16,936 | 19,947 | 19,760 | 23,295 | 21,549 |
| Out-of-pocket | 1,465 | 1,480 | 1,572 | 1,721 | 1,604 | 1,760 |
| **P for trend** |  |  |  |  |  |  |
| VR-12 PCS | 0.287 |  |  |  |  |  |
| VR-12 MCS | 0.473 |  |  |  |  |  |

No significant linear trend was detected for either component (PCS P=0.287, MCS P=0.473). Point estimates were stable across all six years, including the 2020 pandemic year.

**Table 4.**
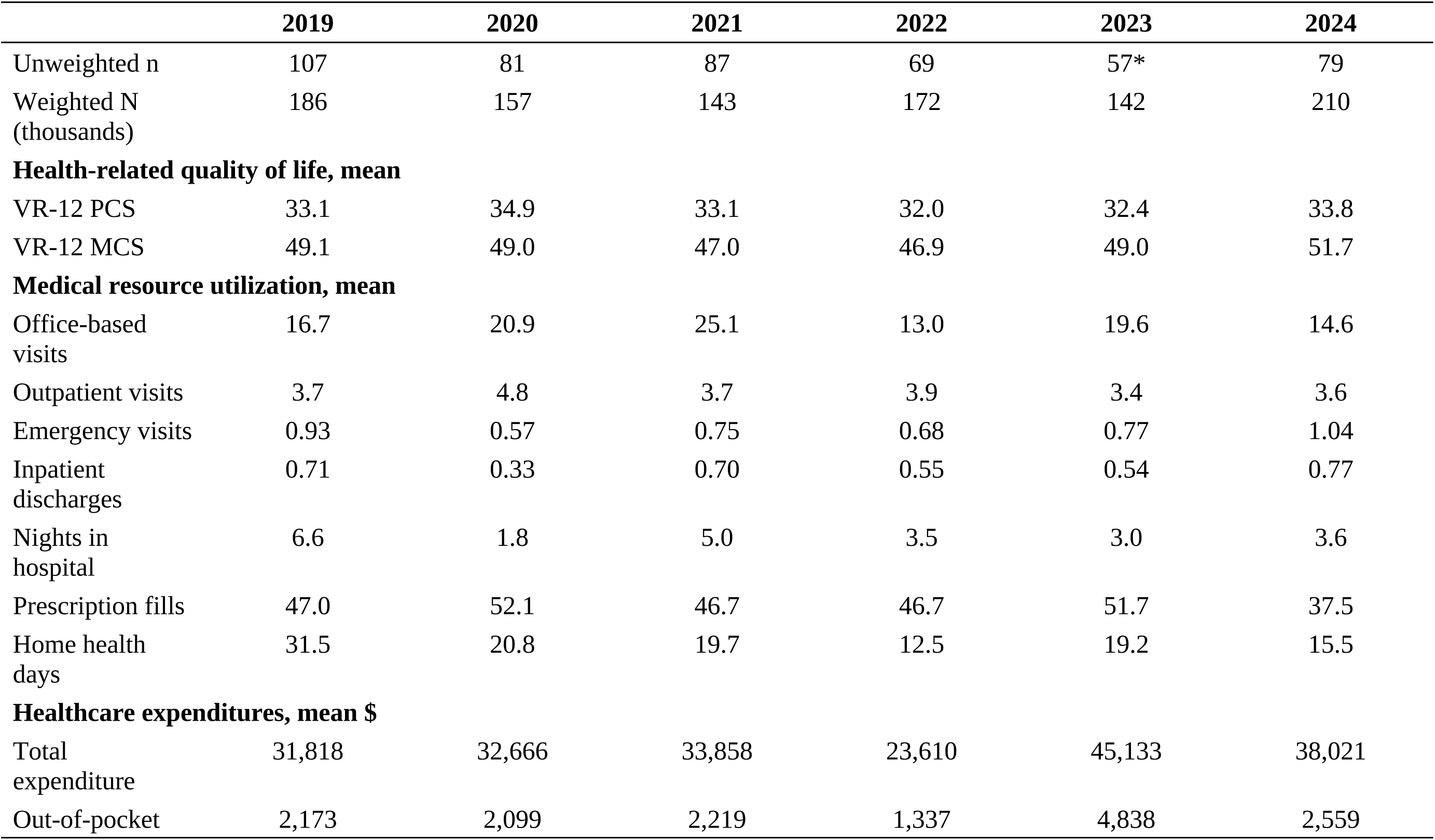
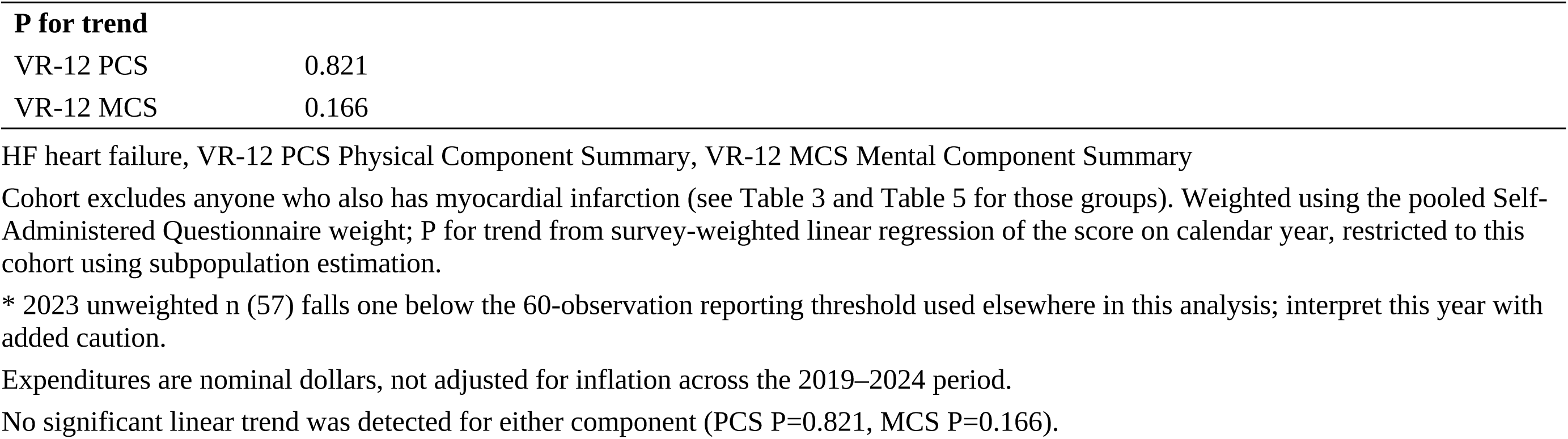
Health-related quality of life, medical resource utilization, and medical expenditures among adults with heart failure only, by year.

|  | 2019 | 2020 | 2021 | 2022 | 2023 | 2024 |
| --- | --- | --- | --- | --- | --- | --- |
| Unweighted n | 107 | 81 | 87 | 69 | 57* | 79 |
| Weighted N<br>(thousands) | 186 | 157 | 143 | 172 | 142 | 210 |
| **Health-related quality of life, mean** |  |  |  |  |  |  |
| VR-12 PCS | 33.1 | 34.9 | 33.1 | 32.0 | 32.4 | 33.8 |
| VR-12 MCS | 49.1 | 49.0 | 47.0 | 46.9 | 49.0 | 51.7 |
| **Medical resource utilization, mean** |  |  |  |  |  |  |
| Office-based<br>visits | 16.7 | 20.9 | 25.1 | 13.0 | 19.6 | 14.6 |
| Outpatient visits | 3.7 | 4.8 | 3.7 | 3.9 | 3.4 | 3.6 |
| Emergency visits | 0.93 | 0.57 | 0.75 | 0.68 | 0.77 | 1.04 |
| Inpatient<br>discharges | 0.71 | 0.33 | 0.70 | 0.55 | 0.54 | 0.77 |
| Nights in<br>hospital | 6.6 | 1.8 | 5.0 | 3.5 | 3.0 | 3.6 |
| Prescription fills | 47.0 | 52.1 | 46.7 | 46.7 | 51.7 | 37.5 |
| Home health<br>days | 31.5 | 20.8 | 19.7 | 12.5 | 19.2 | 15.5 |
| **Healthcare expenditures, mean \$** | | | | | | |
| Total<br>expenditure | 31,818 | 32,666 | 33,858 | 23,610 | 45,133 | 38,021 |
| Out-of-pocket | 2,173 | 2,099 | 2,219 | 1,337 | 4,838 | 2,559 |
| **P for trend** |  |  |  |  |  |  |
| VR-12 PCS | 0.821 |  |  |  |  |  |
| VR-12 MCS | 0.166 |  |  |  |  |  |

\* 2023 unweighted n (57) falls one below the 60-observation reporting threshold used elsewhere in this analysis; interpret this year with added caution.
Expenditures are nominal dollars, not adjusted for inflation across the 2019–2024 period.
No significant linear trend was detected for either component (PCS P=0.821, MCS P=0.166).

**Table 5.**
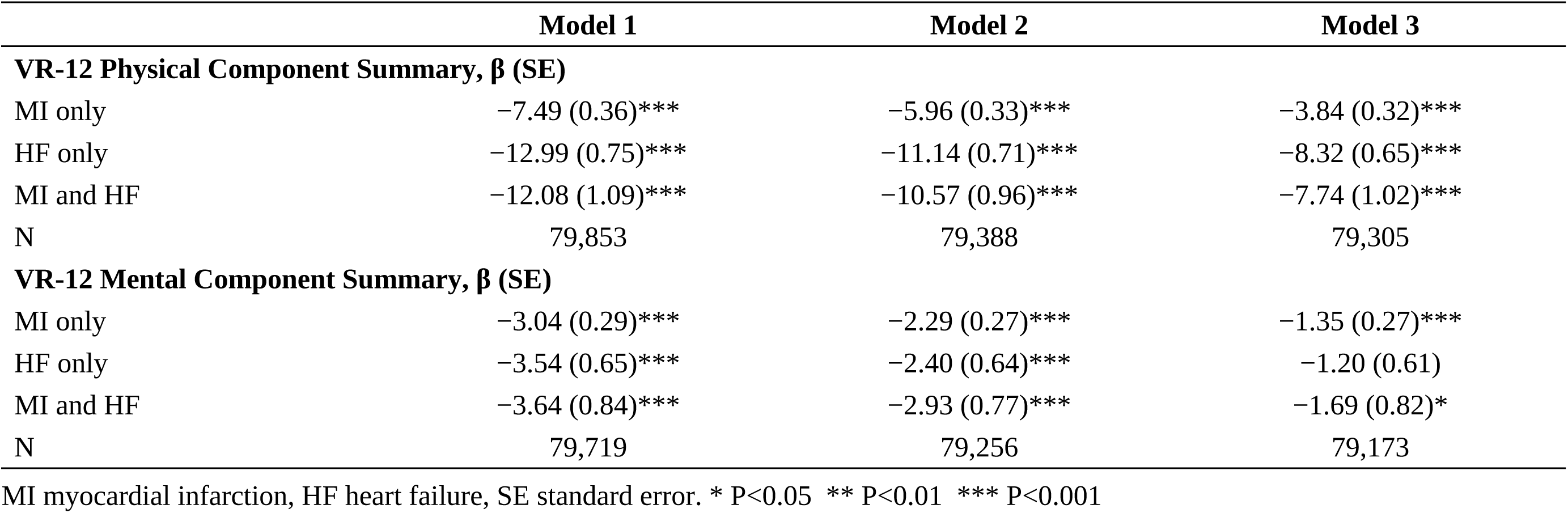
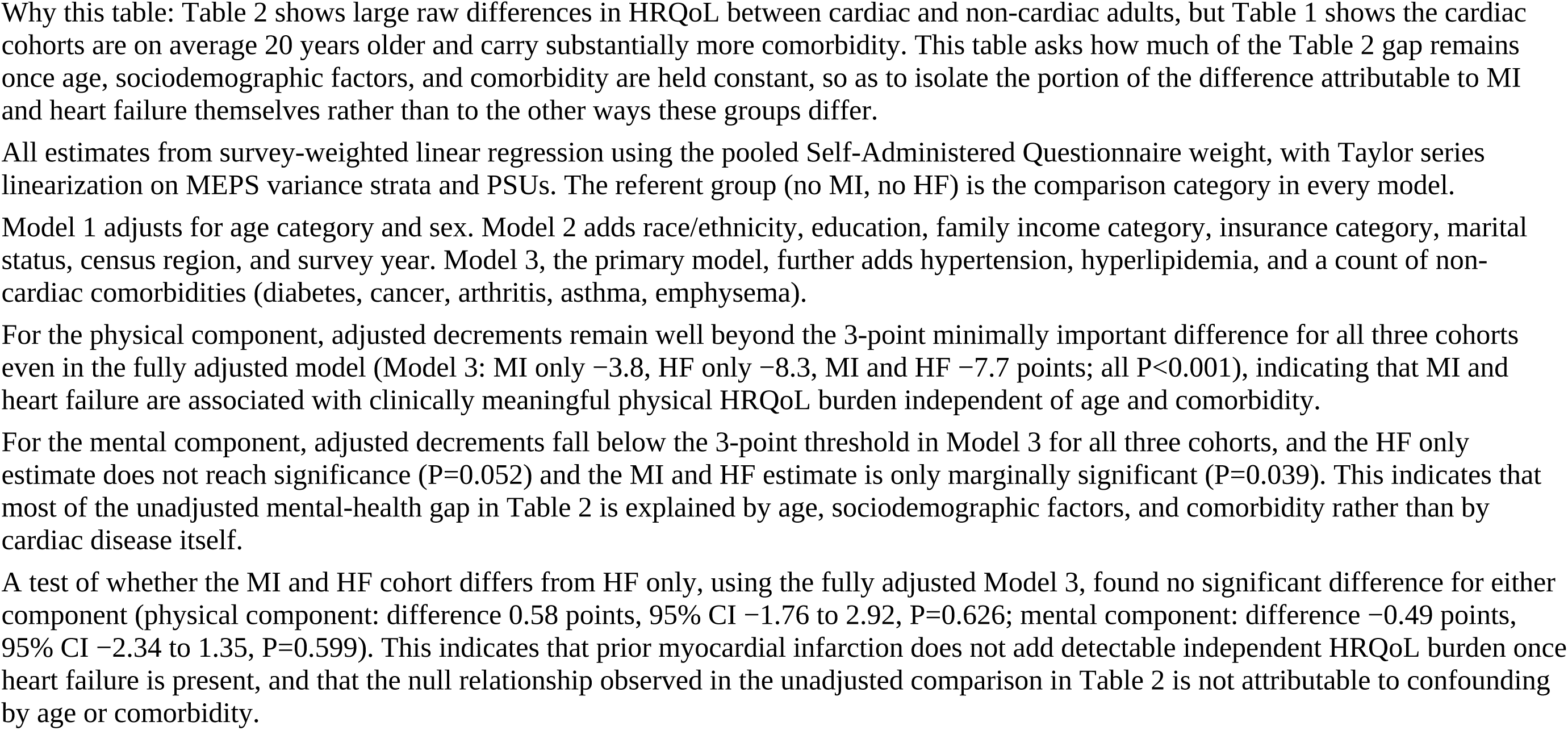
Adjusted differences in health-related quality of life versus adults with neither myocardial infarction nor heart failure.

## DISCUSSION

Overall, this analysis shows that physical quality of life falls steadily across the cardiac cohorts, with heart failure associated with more than twice the difference seen after myocardial infarction. Unadjusted differences from patients with neither condition were 11.4 points in patients with myocardial infarction alone, 17.2 points in patients with heart failure alone, and 16.4 points in patients with both conditions. After adjustment these fell to 3.84 points in patients with myocardial infarction alone, 8.32 points in patients with heart failure alone, and 7.74 points in patients with both conditions, and all three remained beyond the minimally important difference. Differences in the mental component were much smaller. Before adjustment they ranged from 1.6 points in patients with myocardial infarction alone to 2.6 points in patients with both conditions, and after adjustment from 1.20 points in patients with heart failure alone to 1.69 points in patients with both conditions, and none reached the minimally important difference. Patients with both conditions did not differ from patients with heart failure alone on either component. Mean annual expenditure per person was $19,876 in patients with myocardial infarction alone, $34,002 in patients with heart failure alone, and $37,198 in patients with both conditions, against $7,845 in patients with neither condition, and inpatient care was the largest component wherever heart failure was present. The relationship between cardiac diagnosis and quality of life did not change over six years.

This analysis shows that the physical and emotional components of that burden behave very differently. The physical finding fits the clinical character of each condition. Myocardial infarction is an event, and the survey measure captures lifetime history rather than recent illness, so many patients with myocardial infarction alone may be remote from their event and may have recovered functionally. Heart failure is a syndrome defined by exertional limitation, and the physical component summary is built largely from items about walking, climbing stairs, and accomplishing daily tasks. The instrument’s strong response to heart failure and weak response to remote infarction may reflect what the physical component summary is designed to capture.

The absence of any additional burden from prior infarction once heart failure is present is among the more interesting findings. One possible explanation is a ceiling effect. Once a patient is limited enough by heart failure, an additional remote diagnosis may not further restrict the activities the instrument asks about, which are already curtailed. Another possible explanation is survivorship, because patients with both conditions who remain community-dwelling and able to complete a questionnaire may be a selected group. Functional burden in these patients appears to track the presence of heart failure rather than infarct history.

The mental component findings deserve emphasis because they are negative. Before adjustment the difference in mental quality of life approached the minimally important threshold in patients with both conditions, and once age and coexisting illness were accounted for it fell well below that threshold in all 3 cardiac cohorts. This does not mean that patients with cardiac disease are not distressed, because distress screening scores were higher in every cardiac cohort than in patients with neither condition. It suggests instead that distress may track age, income, insurance, and accumulated illness more closely than the cardiac diagnosis itself, and that screening for emotional wellbeing may be better directed by the social and comorbidity profile.

Other findings on expenditure in these conditions are consistent with the pattern reported here. Bhatnagar et al reported total annual expenditure of $28,950 for adults with heart failure against $5,727 without, with hospitalization the largest component at $12,569.^2^ Klein et al reported $21,177 against $5,652 in the same survey and also found inpatient care to be the main driver.^10^ Shah et al found that inpatient costs represented roughly 43% of heart failure spending.^11^ The mental component results align with work in atherosclerotic cardiovascular disease, where adults at high risk for depression reported worse quality of life and higher expenditure than those at low risk.^8^ Karakis et al and Song et al measured other conditions on the same survey; the heart failure physical difference of 8.32 points is roughly twice the 4.0-point difference for epilepsy and larger than the 5.3-point difference for severe asthma.^12,13^ This is a burden that expenditure figures alone do not capture. Further studies may examine the direction of this relationship.

Quality of life in these cohorts did not change between 2019 and 2024 in any of the 4 groups. Point estimates were stable through 2020, the first year of the COVID-19 pandemic. No change was detected over this period, although annual cohort sizes are small and a modest change may not have been detected.

To our knowledge, this is the first study to quantify the quality of life burden of myocardial infarction and heart failure against the general US adult population using a standard instrument. The physical burden in patients with heart failure alone was more than twice that in patients with myocardial infarction alone, and no additional decrement appeared in patients with both conditions beyond patients with heart failure alone. The emotional burden was instead explained largely by age and coexisting illness rather than by the cardiac diagnosis itself. Six pooled years of data allowed the relationship to be examined across a period that included the pandemic, and mutually exclusive cohorts allowed each condition to be estimated without contamination by the other.

These findings need to be interpreted in the context of several limitations. The two conditions are not identified the same way. Myocardial infarction is a lifetime self-reported diagnosis requiring no healthcare contact, while heart failure is recorded only when reported in connection with care received that year, which raises utilization and expenditure in patients with heart failure by construction. Expenditure comparisons are unadjusted and in nominal dollars, and patients with neither condition are on average two decades younger with far less coexisting illness, so the difference between $37,198 in patients with both conditions and $7,845 in patients with neither condition is not an estimate of the cost of cardiac disease. Finally, the cross-sectional design cannot establish cause and effect between cardiac diagnosis and quality of life.

Functional assessment in cardiac clinics may be better prompted by the presence of heart failure than by infarct history, and screening for emotional wellbeing may be better directed by social circumstance and accumulated illness than by the cardiac diagnosis alone. Future studies should repeat this comparison with inflation-adjusted expenditure and adjusted cost models, phenotype heart failure using linked clinical data, and use the two questionnaire administrations within each survey panel to place quality of life before expenditure in time.

## CONCLUSIONS

To summarize, we found that physical quality of life falls progressively across US adults with cardiac disease, with heart failure rather than prior myocardial infarction accounting for most of that decline. Patients with heart failure alone scored more than twice as far below patients with neither condition as patients with myocardial infarction alone, and a history of infarction added no measurable further burden once heart failure was established. Differences in mental quality of life were smaller and did not persist after adjustment for age and coexisting illness. Spending and patient-reported quality of life did not rank these 4 groups in the same order, which suggests that expenditure alone understates what patients with heart failure experience. Clinicians should be aware that functional burden in cardiac disease tracks the presence of heart failure, and that emotional wellbeing in these patients tracks social circumstance and accumulated illness more closely than it tracks the cardiac diagnosis.

## Data Availability

All data is available on request.

## Acknowledgments

None.

## Data Availability

The Medical Expenditure Panel Survey public use files analyzed in this study are available from the Agency for Healthcare Research and Quality at https://meps.ahrq.gov.

## Non-standard Abbreviations and Acronyms

MI: myocardial infarction
HF: heart failure
MEPS: Medical Expenditure Panel Survey
VR-12: Veterans RAND 12-Item Health Survey
PCS: physical component summary
MCS: mental component summary

