## supplemental tables and figures for "Health-Related Quality of Life, Healthcare Utilization, and Expenditure in US Adults With Myocardial Infarction and Heart Failure: The Medical Expenditure Panel Survey 2019 to 2024"

### SUPPLEMENTAL TABLE

**Table S1. Health-related quality of life, medical resource utilization, and medical expenditures among adults with both myocardial infarction and heart failure, by year**

| | 2019 | 2020 | 2021 | 2022 | 2023 | 2024 |
|---|---|---|---|---|---|---|
| Unweighted n | 57 | 52 | 44 | 36 | 23 | 24 |
| Weighted N (thousands) | 108 | 110 | 82 | 97 | 98 | 65 |
| **Health-related quality of life, mean** | | | | | | |
| VR-12 PCS | 30.7 | 35.8 | 35.0 | 30.0 | 39.6 | 33.3 |
| VR-12 MCS | 51.0 | 50.0 | 47.6 | 48.9 | 48.8 | 47.9 |
| **Medical resource utilization, mean** | | | | | | |
| Office-based visits | 17.9 | 16.6 | 22.4 | 17.9 | 13.1 | 14.1 |
| Outpatient visits | 2.9 | 3.0 | 3.4 | 4.5 | 5.2 | 3.8 |
| Emergency visits | 1.09 | 0.86 | 0.72 | 0.89 | 1.32 | 1.25 |
| Inpatient discharges | 0.68 | 0.67 | 0.56 | 0.92 | 1.03 | 0.76 |
| Nights in hospital | 4.9 | 5.3 | 2.8 | 7.7 | 4.2 | 2.9 |
| Prescription fills | 45.7 | 36.4 | 46.5 | 47.6 | 38.8 | 42.6 |

| | 2019 | 2020 | 2021 | 2022 | 2023 | 2024 |
|---|---|---|---|---|---|---|
| Home health days | 30.5 | 13.3 | 21.4 | 40.4† | 8.7 | 10.5 |
| **Healthcare expenditures, mean $** | | | | | | |
| Total expenditure | 29,598 | 37,853 | 24,648 | 41,510 | 47,562 | 42,551 |
| Out-of-pocket | 1,415 | 3,856 | 1,844 | 2,034 | 3,018 | 12,476† |
| **P for trend (descriptive only, see note)** | | | | | | |
| VR-12 PCS | 0.242 | | | | | |
| VR-12 MCS | 0.262 | | | | | |

VR-12 PCS Physical Component Summary, VR-12 MCS Mental Component Summary

This cohort has both myocardial infarction and heart failure (see Table 3 and Table 4 for the exclusive groups). Unweighted cell sizes range from 23 to 57 per year.

† These cells are driven by a small number of high-cost or high-utilization respondents and should be interpreted with caution; the corresponding 95% confidence intervals are wide and in some cases include zero.

The trend test is shown for completeness only. At 23-57 observations per year this analysis is underpowered to detect a linear trend and the result should not be interpreted as evidence of stability or change; it is reported descriptively rather than as a formal finding.

Expenditures are nominal dollars, not adjusted for inflation across the 2019–2024 period.

Across all three by-year tables (3, 4, and 5) and the pooled six-year estimates in Table 2, no cohort showed a statistically significant trend in VR-12 physical or mental component scores over 2019-2024, a period spanning the COVID-19 pandemic.

### SUPPLEMENTAL FIGURES

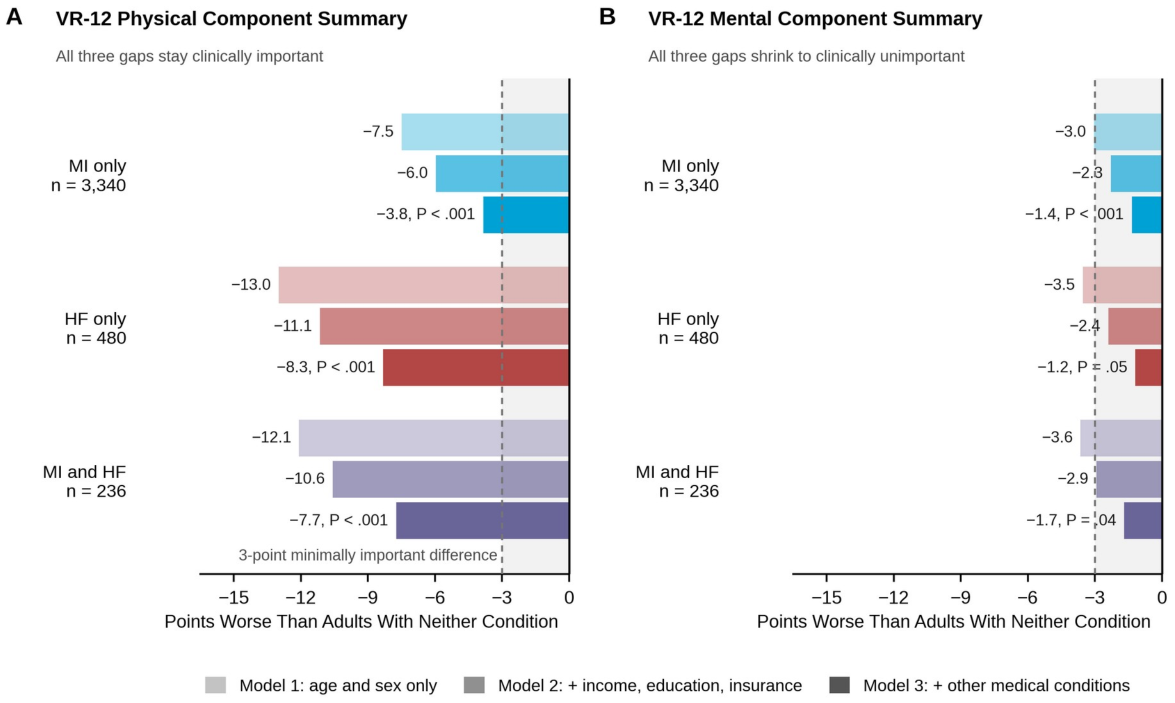


Each cohort has three bars, one per adjustment model, all comparing that cohort with adults having neither condition. Longer bars mean a larger quality of life gap. Bars shorten from Model 1 to Model 3 because progressively more of the raw gap is explained by age, social circumstances, and other illnesses rather than by cardiac disease itself. Only the mental component bars finish inside the shaded zone, where a difference is too small for a patient to notice. P values are shown for the fully adjusted Model 3 only.
HF indicates heart failure; MI, myocardial infarction.

**Figure S1. Adjusted differences under three nested models.** A, Physical Component Summary. B, Mental Component Summary. Longer bars indicate a larger difference. Model 1 adjusts for age and sex, Model 2 adds sociodemographic covariates, and Model 3 adds coexisting conditions. P values are shown for the fully adjusted Model 3 only.

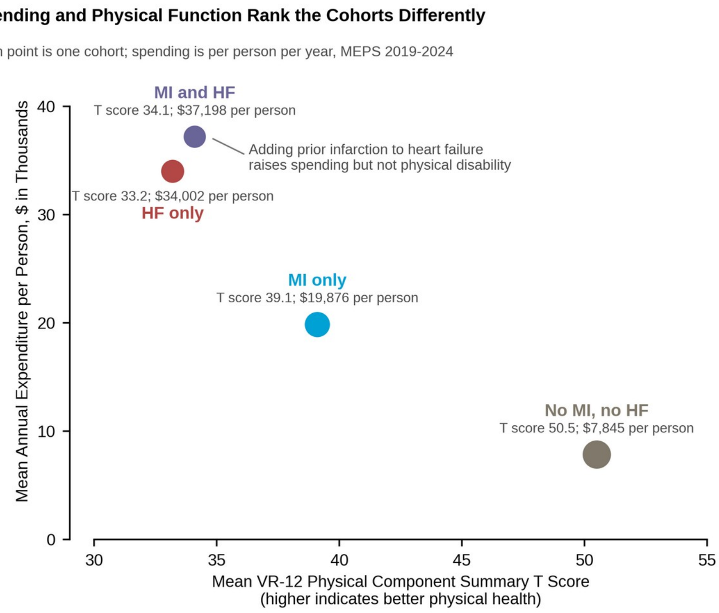


Marker area scales with the logarithm of unweighted sample size. Both axes show unadjusted weighted means, and expenditures are in nominal dollars per person per year. The cohort that spends the most is not the cohort with the worst physical function. HF indicates heart failure; MI, myocardial infarction.

**Figure S2. Physical component score against total expenditure by cohort.** Each point is one cohort. Marker area scales with the logarithm of unweighted sample size. Both axes show unadjusted weighted means.

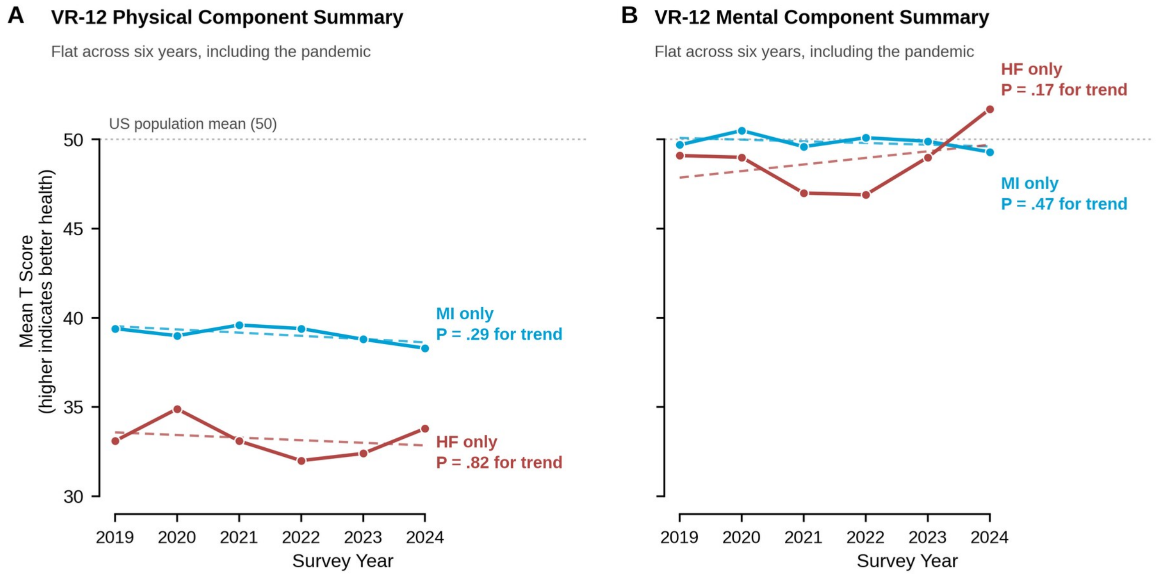


Solid lines connect annual weighted means; dashed lines are the fitted linear trend, shown to make the absence of change visible. Neither cohort changed significantly on either component. The small group with both conditions is omitted here because it rests on 23 to 57 observations per year; it appears in the supplement. HF indicates heart failure; MI, myocardial infarction.

**Figure S3. Component summary scores by calendar year.** A, Physical Component Summary. B, Mental Component Summary. Solid lines connect annual weighted means and dashed lines are the fitted linear trend. The cohort with both conditions is omitted because it rests on 23 to 57 observations per year.
